# SARS-CoV-2 antibody binding and neutralization in dried blood spot eluates and paired plasma

**DOI:** 10.1101/2021.08.02.21261504

**Authors:** Hannah L. Itell, Haidyn Weight, Carolyn S. Fish, Jennifer K. Logue, Nicholas Franko, Caitlin R. Wolf, Denise J. McCulloch, Jared Galloway, Frederick A. Matsen, Helen Y. Chu, Julie Overbaugh

## Abstract

Widescale assessment of SARS-CoV-2-specific antibodies is critical to understanding population seroprevalence, correlates of protection, and the longevity of vaccine-elicited responses. Most SARS-CoV-2 studies characterize antibody responses in plasma/sera. While reliable and broadly used, these samples pose several logistical restrictions such as requiring venipuncture for collection and cold chain for transportation and storage. Dried blood spots (DBS) overcome these barriers as they can be self-collected by fingerstick and mailed and stored at ambient temperature. Here, we evaluate the suitability of DBS for SARS-CoV-2 antibody assays by comparing several antibody responses between paired plasma and DBS from SARS-CoV-2 convalescent and vaccinated individuals. We found that DBS not only reflected plasma antibody binding by ELISA and epitope profiles using phage-display, but also yielded SARS-CoV-2 neutralization titers that highly correlated with paired plasma. Neutralization measurement was further streamlined by adapting assays to a high-throughput 384-well format. This study supports the adoption of DBS for numerous SARS-CoV-2 binding and neutralization assays.

## INTRODUCTION

The ability to detect and characterize antibodies targeting SARS-CoV-2 proteins in plasma has been one of the most informative tools during the ongoing COVID-19 pandemic. Serological testing identifies cases of previous infection, including those not revealed by symptoms and viral testing, and therefore provides a more accurate estimate of regional exposure rates than cumulative RT-PCR-based testing (1). In addition to public health surveillance, assessing SARS-CoV-2 antibody binding and functional activity has provided insight on the magnitude, features, and durability of antibody responses elicited by natural infection (2-4) and, more recently, vaccination (5-7). Antibody-based investigations have therefore informed lockdown regulations, vaccine development (8-11), possible correlates of protection (4, 12, 13), and more.

Though tremendous progress has been made to curb the pandemic, several pressing questions remain that necessitate the continued evaluation of SARS-CoV-2 antibodies in large cohorts around the world. Namely, it is unknown how long immunity conferred by infection or vaccination lasts and whether these forms of immunity are protective against viral variants. Addressing these questions will likely require global sample collection for years to come, as immunity against variants must be investigated as they emerge and there is an ever-growing list of vaccines (8-11, 14), second dose combinations (15), and boosters (16) that must be evaluated for longevity of protection.

The enormity of this task underscores the need to optimize the feasibility and practicality of SARS-CoV-2 antibody investigations. Some of the most expensive and time-consuming aspects of these studies involve the collection, shipping, and storage of plasma and sera samples. These sample types are collected via venipuncture by trained phlebotomists typically in the clinic and are immediately stored in a refrigerator or freezer and longer-term at -20ºC. The cold chain must be maintained during transportation for biospecimen integrity. Therefore, if individuals cannot access a clinic, phlebotomists are unavailable or over-burdened, the cold chain is not maintained, or long-term -20ºC storage is too costly, using plasma and/or sera sampling becomes logistically prohibitive.

Dried blood spots (DBS) have previously been used for nucleic acid and antibody testing and represent a more practical sampling type for SARS-CoV-2 antibody studies. DBS cards are prepared by spotting whole blood onto filter paper cards. Once dried, cards are stable at ambient temperature for at least a few weeks (17, 18) and can be mailed without cold chain or special authorizations for international shipments (19). DBS cards can therefore be collected at home via a non-invasive, self-administered fingerstick and mailed to the laboratory, where small DBS discs are excised and eluted for assay use (20). DBS cards bypass the need of trained personnel for collection or the cold chain for shipping, which not only lowers costs and reduces frontline worker demand, but also facilitates sampling hard-to-reach populations.

The utility and reliability of DBS cards have been demonstrated for several decades. DBS sampling has been used for infant metabolic screening since the 1960s (21, 22) and now has broad applications with over 2,000 analytes measured in DBS eluates to date (23). In the context of infectious disease research, DBS cards are valuable for the surveillance and study of HIV and tropical diseases (24-26) by enabling sampling in resource-limited areas. Therefore, when the current pandemic began, several groups sought to verify the use of DBS eluates for SARS-CoV-2 antibody assays. The most robust of these studies compared antibody binding measurements between paired DBS eluates and plasma or sera from COVID-19 convalescent individuals (27-36) and, impressively, all have found strong agreement between sample types. Despite this supportive evidence, the majority of these reports did not use self-collected, mailed-in fingerstick DBS cards and the compatibility of DBS with other assay formats, including neutralization, has not been carefully examined for SARS-CoV-2.

In this study, we addressed gaps in the SARS-CoV-2 DBS field by collecting paired plasma and DBS cards from COVID-19 convalescent and SARS-CoV-2 vaccinated individuals. We assessed the agreement between sample types for several antibody-based methods, including a widely used receptor-binding domain (RBD) ELISA (37, 38), a comprehensive phage-display approach (39), and a SARS-CoV-2 spike neutralization assay (40), which we optimized here for higher throughput. For all approaches, we found consistently high agreement between sample types, including between paired plasma and eluates from self-collected fingerstick DBS cards. Additionally, we evaluated the stability of antibodies on DBS cards after six months at room temperature and found no substantial decline in SARS-CoV-2-specific IgG binding. These results support the adoption of DBS sampling for SARS-CoV-2 antibody studies as a reliable, feasible addition to current plasma and sera approaches.

## RESULTS

### Characteristics of paired sample groups

Individuals with previous COVID-19 infection and/or SARS-CoV-2 vaccination were enrolled into a prospective observational study at the University of Washington Seattle, Washington, United States, and paired plasma and DBS cards were collected from these individuals. DBS cards were self-prepared at home via fingerstick (FS DBS) or at the clinic by spotting venous blood onto cards immediately after venipuncture (VB DBS). Paired samples were designated into one of three sample groups depending on DBS type and history of infection and/or vaccination, as outlined in **Figure 1**.

**Figure 1.**
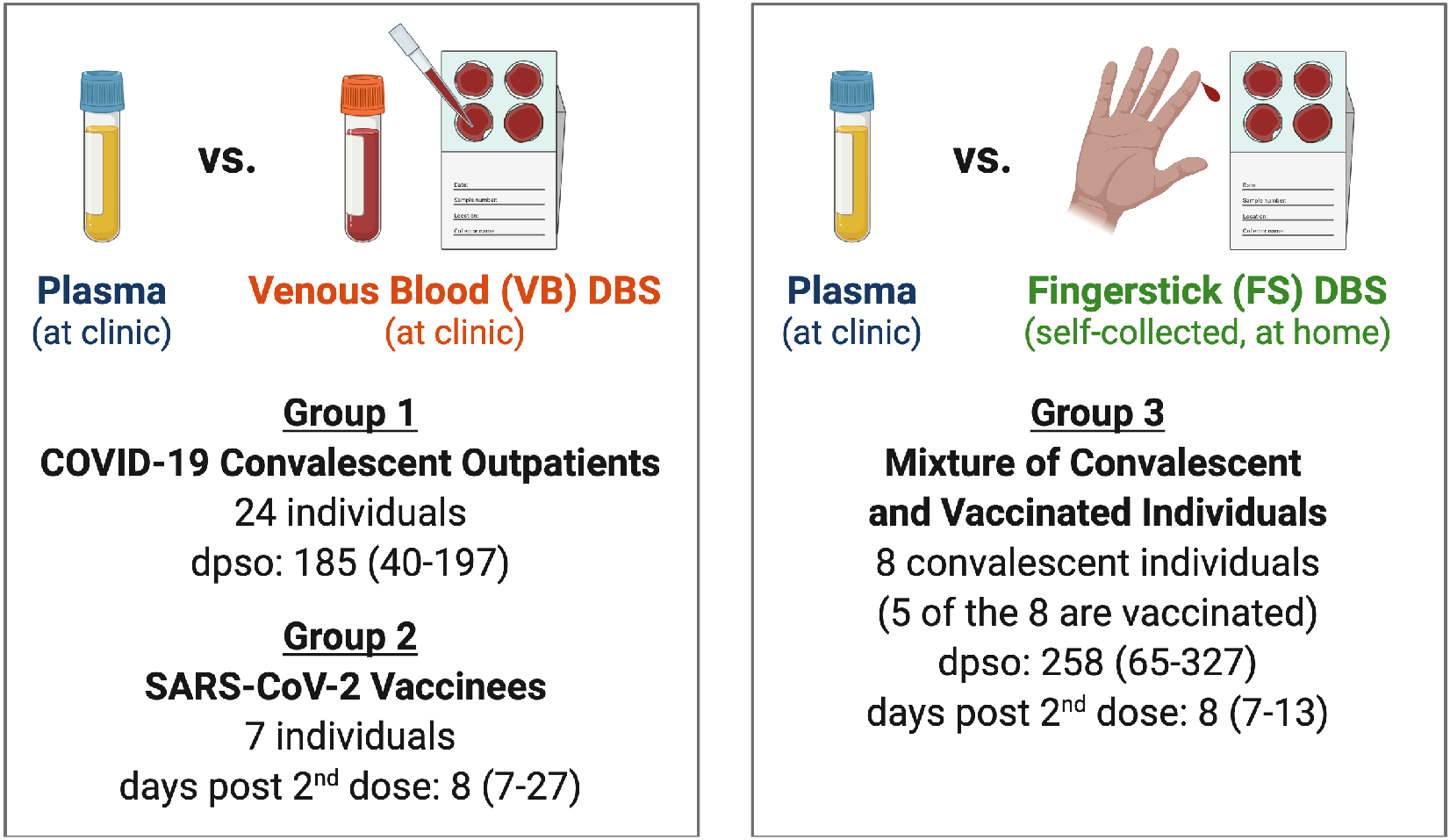
Characteristics of paired sample groups. Paired samples were assigned to one of three study groups based on the sample types collected and participant SARS-CoV-2 infection and vaccination history as outlined in the figure. All vaccinated individuals received two doses of the Moderna mRNA-1273 or Pfizer/BioNTech BNT162b2 vaccines. Median (range). DPSO: days post symptom onset. Created with BioRender.com.

### VB DBS eluates reflect total and SARS-CoV-2-specific IgG levels in paired plasma from COVID-19 convalescent patients

We first measured total IgG levels in Group 1 VB DBS eluates and paired plasma (n=24) to estimate the volume of plasma on each 6-mm DBS subpunch. IgG concentrations determined by ELISA ranged from 110 to 317 µg/mL in VB DBS samples eluted in 100 µL of PBS-Tween (**Figure 2A**), which indicates that a median of 17.9 µg IgG was eluted from each disc. By comparing the IgG amount on each VB DBS disc to the levels in paired plasma, we calculated that each 6-mm disc from Group 1 contains a median of 5.6 µL of plasma (range: 4.2-7.9). Therefore, there is an approximate 20-fold initial plasma dilution introduced when DBS discs are eluted that must be accounted for in downstream assays. Overall, there was a strong correlation in IgG levels measured between these two sample types (Pearson R: 0.83; **Figure 2A**).

**Figure 2.**
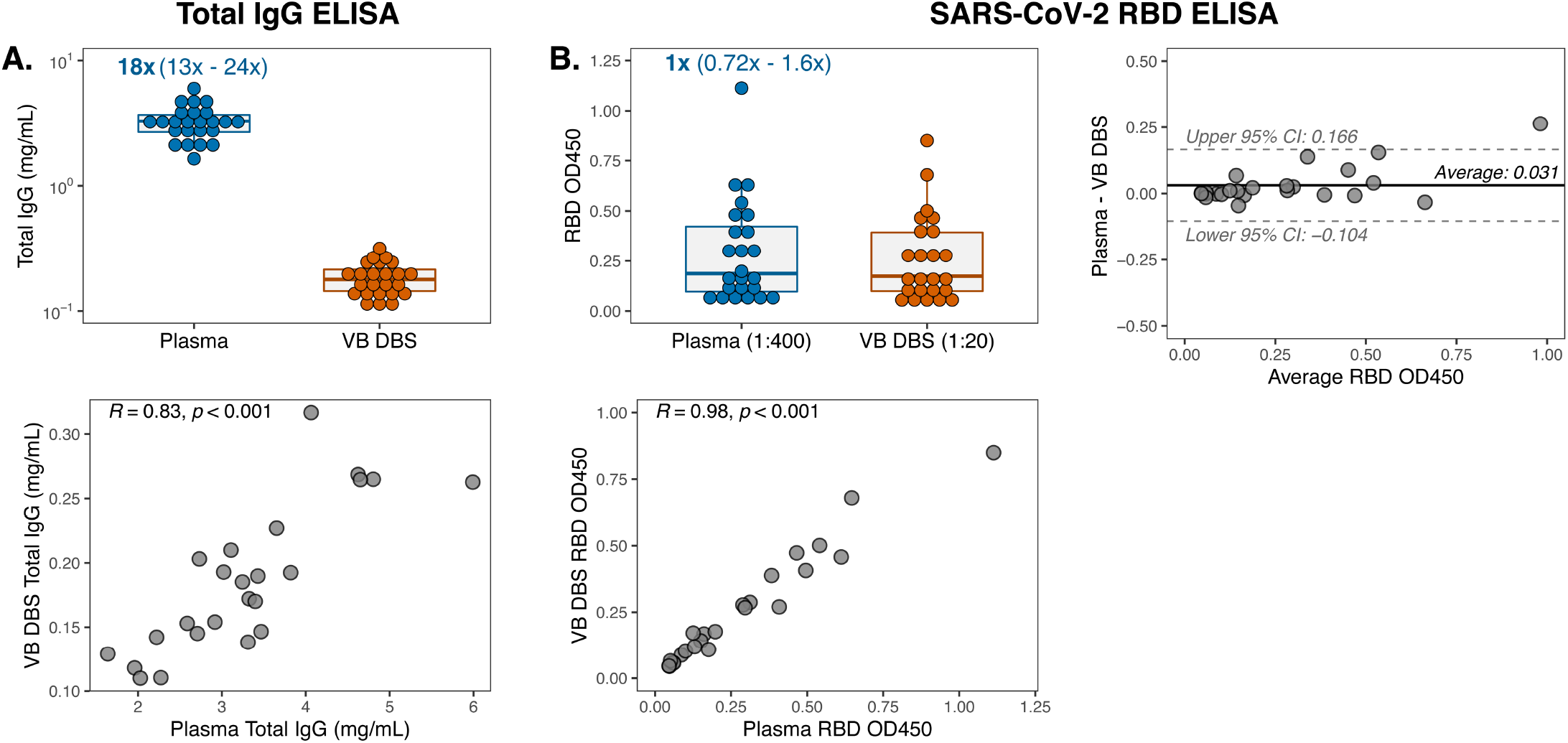
VB DBS eluates reflect total and SARS-CoV-2-specific IgG levels in paired plasma from COVID-19 convalescent patients. (**A**) Total IgG concentrations in Group 1 VB DBS eluates and paired plasma (n=24 pairs) and the Pearson R correlation comparing IgG levels between sample types. The median and range fold differences between paired samples are reported above the plasma data. (**B**) IgG reactivity to SARS-CoV-2 RBD protein at single dilutions, as indicated. Results were also depicted as a Pearson R correlation and a Bland-Altman plot to compare VB DBS eluate and plasma RBD OD_450_ measurements.

To determine whether VB DBS-eluted antibodies recapitulate plasma SARS-CoV-2 RBD binding, we assayed samples via a widely used in-house RBD ELISA (37, 38). At point dilutions that accounted for the initial ∼20-fold dilution introduced during DBS preparation, paired samples demonstrated very similar RBD OD measurements (median fold change of 1; **Figure 2B**). Similarly, the trends in binding magnitude across individuals strongly agreed between sample types (Pearson R: 0.98; **Figure 2B**). These results demonstrate the sensitivity of DBS eluates to detect antibody binding despite the dilution that occurs due to sample processing. We further assessed eluate and plasma agreement by performing Bland-Altman analysis on the absolute difference between the OD measurements for each sample type (**Figure 2B**). We observed an average bias of +0.031 OD in plasma (95% confidence interval (CI): -0.104-0.166) and only one sample pair, which also had the highest average OD, exceeded the 95% CIs. This average bias falls below the typical level of background observed in empty RBD ELISA wells (0.05 OD, data not shown). Therefore, we found high concordance in SARS-CoV-2 RBD binding via a standard ELISA between paired VB DBS eluates and plasma from convalescent individuals.

### Polyclonal antibody response characteristics and epitope specificities can be defined using DBS

We next sought to examine the utility of DBS to capture epitope specificity and to determine whether antibodies to the dominant SARS-CoV-2 epitopes present in polyclonal plasma are preserved in VB DBS eluates. To address this, we tested 22 sample pairs from Group 1 for binding to a library of phage displaying SARS-CoV-2 peptides. This phage library expresses 39 amino acid-long peptides that span the entire SARS-CoV-2 proteome in 20 amino acid increments, for a total of 480 SARS-CoV-2 peptides (39). Ten micrograms of IgG for each sample was incubated with the peptide library. Antibody-phage complexes were then immunoprecipitated and sequenced to define the linear peptides that antibodies bound to in the two sample types.

In agreement with a previous report from our group on convalescent plasma (39), the proteins with the highest magnitude of peptide binding as measured by counts per million (CPM) were ORF1ab, nucleocapsid, and spike for both sample types. Within these dominant sites, VB DBS eluates closely recapitulated the binding profiles observed in plasma on a cohort-wide scale (**Figure 3A**). Moreover, average CPMs for each SARS-CoV-2 peptide across the 22 individuals strongly correlated between sample types (Pearson R: 0.87; **Figure 3B**), indicating cohort-wide agreement across all proteins. To contextualize the magnitude of this correlation, we compared average CPMs across the 22 plasma samples between duplicate assay wells and observed a very similar level of agreement (Pearson R: 0.94; data not shown), suggesting that antibodies eluted from DBS cards recapitulate cohort-wide plasma epitope profiling results nearly to the level of within-assay plasma replicates.

**Figure 3.**
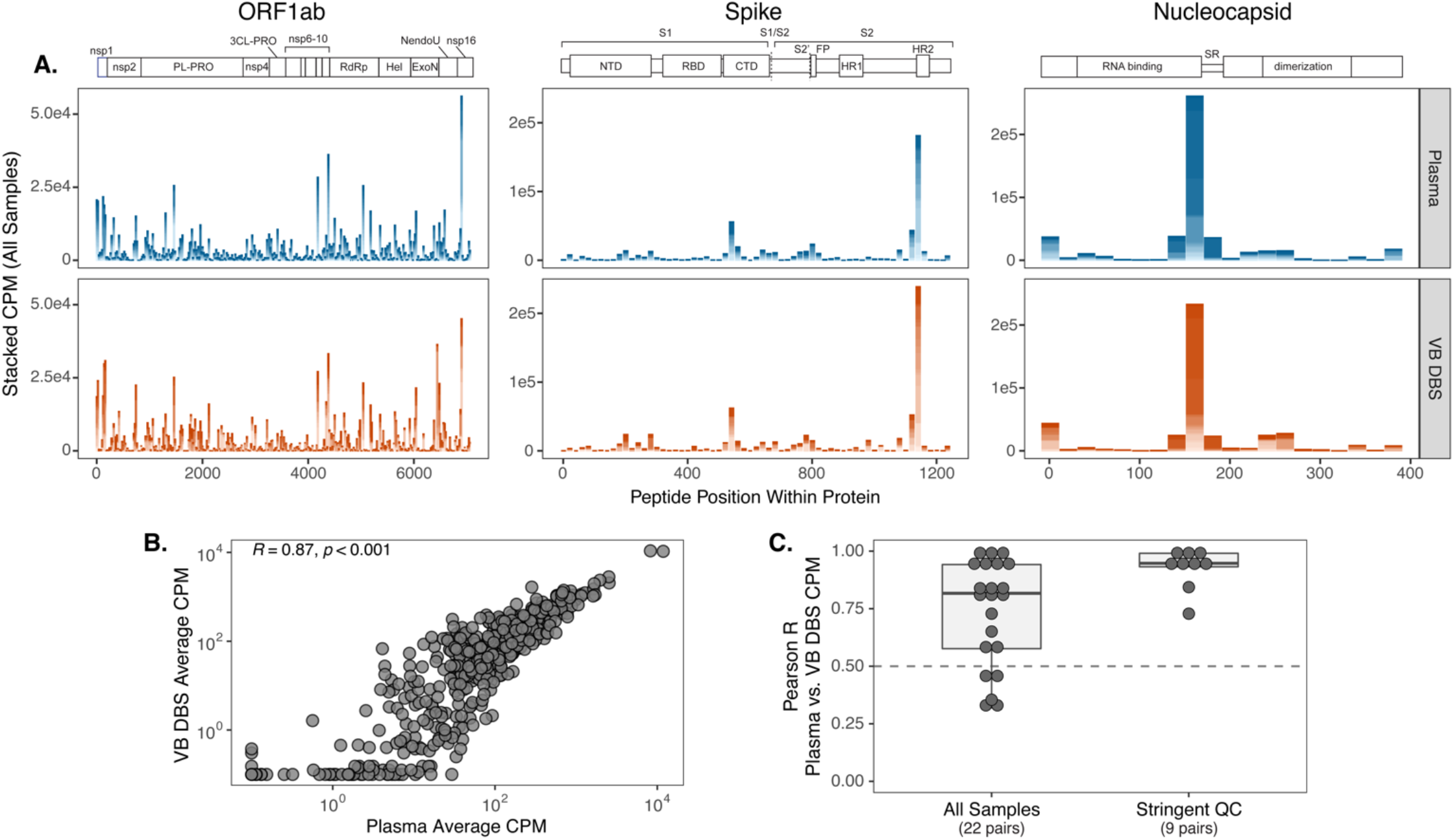
Polyclonal antibody response characteristics and epitope specificities can be defined using DBS. (**A**) Counts per million (CPM) of SARS-CoV-2 peptides within the three proteins with the highest magnitude of binding: ORF1ab, spike, and nucleocapsid. Peptide CPMs are stacked across Group 1 individuals for each sample type (n=22). (**B**) Pearson R correlation of average peptide CPMs between sample types. (**C**) CPM values between sample pairs were correlated for each individual and the distribution of correlation coefficients is shown for all sample pairs and those with high replicate reproducibility (QC: quality control).

We also compared epitope profiles between VB DBS eluates and plasma on the individual level by correlating SARS-CoV-2 peptide CPM results between sample pairs. We observed strong agreement between sample pairs with a median Pearson R coefficient of 0.82 and 17/22 pairs having Pearson R values greater than 0.5 (**Figure 3C**). To evaluate whether sample data quality influences agreement between paired samples, we correlated SARS-CoV-2 peptide CPM values between duplicate assay wells and focused on those sample pairs with high DBS replicate agreement (replicate Pearson R>0.5; n=9 pairs). Peptide counts between the resulting plasma and VB DBS pairs were consistently highly correlated (median Pearson R: 0.95, **Figure 3C**), which suggests that DBS eluates more closely reflect paired plasma results when DBS replicate reproducibility is high. These data support the use of VB DBS eluates in place of plasma for both cohort-wide epitope mapping investigations and for the evaluation of individual binding profiles.

### Neutralizing antibody levels in vaccinated individuals can be accurately measured using DBS

Recent studies continue to support antibody-mediated neutralization of SARS-CoV-2 as an important immune response conferring protection against infection and COVID-19 disease (12, 13, 41). Therefore, there is a continued need to evaluate levels of neutralization in large cohorts not only to understand the duration of this response after vaccination but also to assess neutralization efficacy against emerging viral variants. To determine whether DBS eluates capture plasma neutralization activity, we collected paired plasma and VB DBS cards from seven individuals at a median of eight days after their second Moderna mRNA-1273 or Pfizer/BioNTech BNT162b2 SARS-CoV-2 vaccination (Group 2; **Figure 1**). For this experiment, DBS discs were eluted in 50 µL serum-free media instead of 100 µL PBS-Tween to concentrate the eluate and ensure its compatibility with cell culture (see Methods). We first measured total IgG levels in these concentrated, media-eluted VB DBS eluates and paired plasma and observed a median 8-fold difference between sample types (**Figure 4A**). This was approximately half of that observed with VB DBS eluates eluted in twice in the volume (**Figure 2A**) and corresponded with a median of 6.3 µL plasma per 6-mm disc, which was very similar to the prediction from the larger elution volume. Consistent with the total IgG results with Group 1 samples, Group 2 VB DBS eluates and plasma IgG levels strongly correlated (Pearson R=0.91, **Figure 4A**).

**Figure 4.**
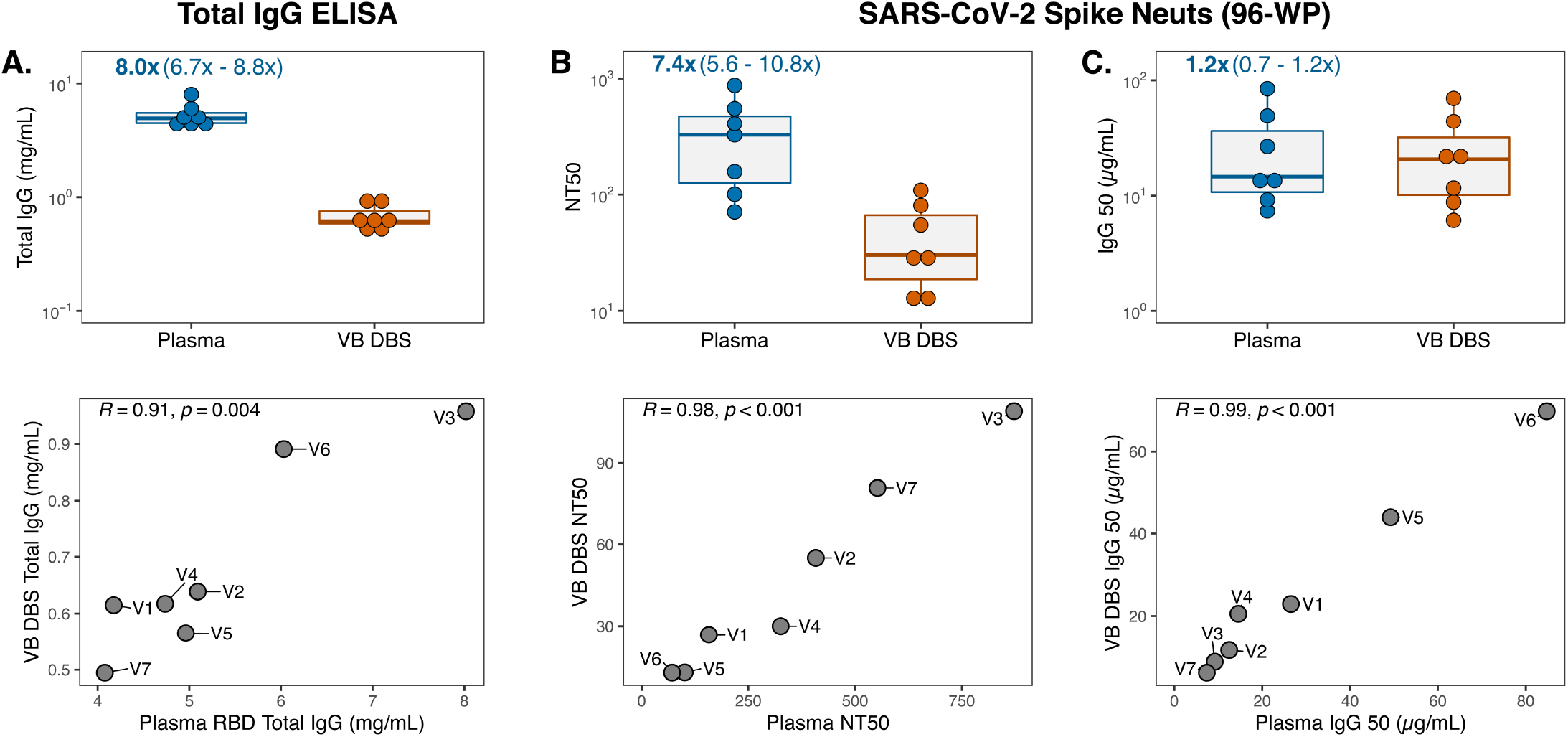
Neutralizing antibody levels in vaccinated individuals can be accurately measured using DBS. (**A**) Total IgG levels in concentrated, media-eluted Group 2 VB DBS eluates and paired plasma (n=7 pairs) and respective Pearson R correlations between sample types. Data points in the correlation plot are labeled with Group 2 participant IDs (V1-V7). (**B**) Neutralization titer 50 (NT) and (**C**) IgG_50_ (total IgG concentration divided by NT_50_) results for sample type pairs and Pearson R correlations.

To evaluate neutralization activity in plasma and DBS eluates, we leveraged a previously described SARS-CoV-2 spike-pseudotyped lentiviral particle assay (40) and found that all plasma samples from vaccinated individuals had detectable neutralization titers (NT_50_ range: 71.1-872.1). VB DBS eluates demonstrated neutralization titers 7.4-fold lower than plasma (**Figure 4B**), which is consistent with the 8-fold difference in total IgG levels (**Figure 4A**). Remarkably, the NT_50_ values between sample types correlated very strongly (**Figure 4B**, Pearson R=0.98). To understand whether the 7.4-fold difference in NT_50_ values between sample types was driven by differences in total IgG content, we calculated the IgG_50_ for each sample (IgG_50_=IgG concentration/NT_50_). After this normalization, there was no significant difference in neutralization activity between sample types (**Figure 4C**, Wilcoxon Rank p-value=0.22, fold difference=1.2) and the strong correlation between sample pairs was maintained (**Figure 4C**, Pearson R=0.99). Importantly, these results were not impacted by background signal from either sample type, as a pre-pandemic sera pool and an eluate from a blank DBS card did not yield any neutralization activity (**Figure S1**).

These findings indicate that antibodies eluted from DBS cards recapitulate plasma NT_50_ trends and IgG_50_ magnitude. However, a drawback of the spike neutralization assay is that it requires large sample volumes due to its 96-well plate (WP) layout, which could be limiting in the case of DBS sampling. We therefore adapted the existing SARS-CoV-2 spike neutralization assay to a high-throughput 384-WP format, which uses a third of the sample volume and assays five times the number of samples per plate. To compare assay formats, we re-assayed the same seven sample pairs and found that NT_50_ measurements strongly correlated between plates for both sample types, which demonstrates that trends in neutralization titers across samples and sample types are maintained in the 384-WP format (**Figure 5A**, Pearson R=0.95). Additionally, as we observed in the 96-WP assay, NT_50_ and IgG_50_ values were strongly correlated between sample types in the high-throughput format (**Figure 5B-C**). The 384-WP spike neutralization assay is thus a suitable and more practical alternative to the traditional 96-WP format for both DBS eluates and plasma.

**Figure 5.**
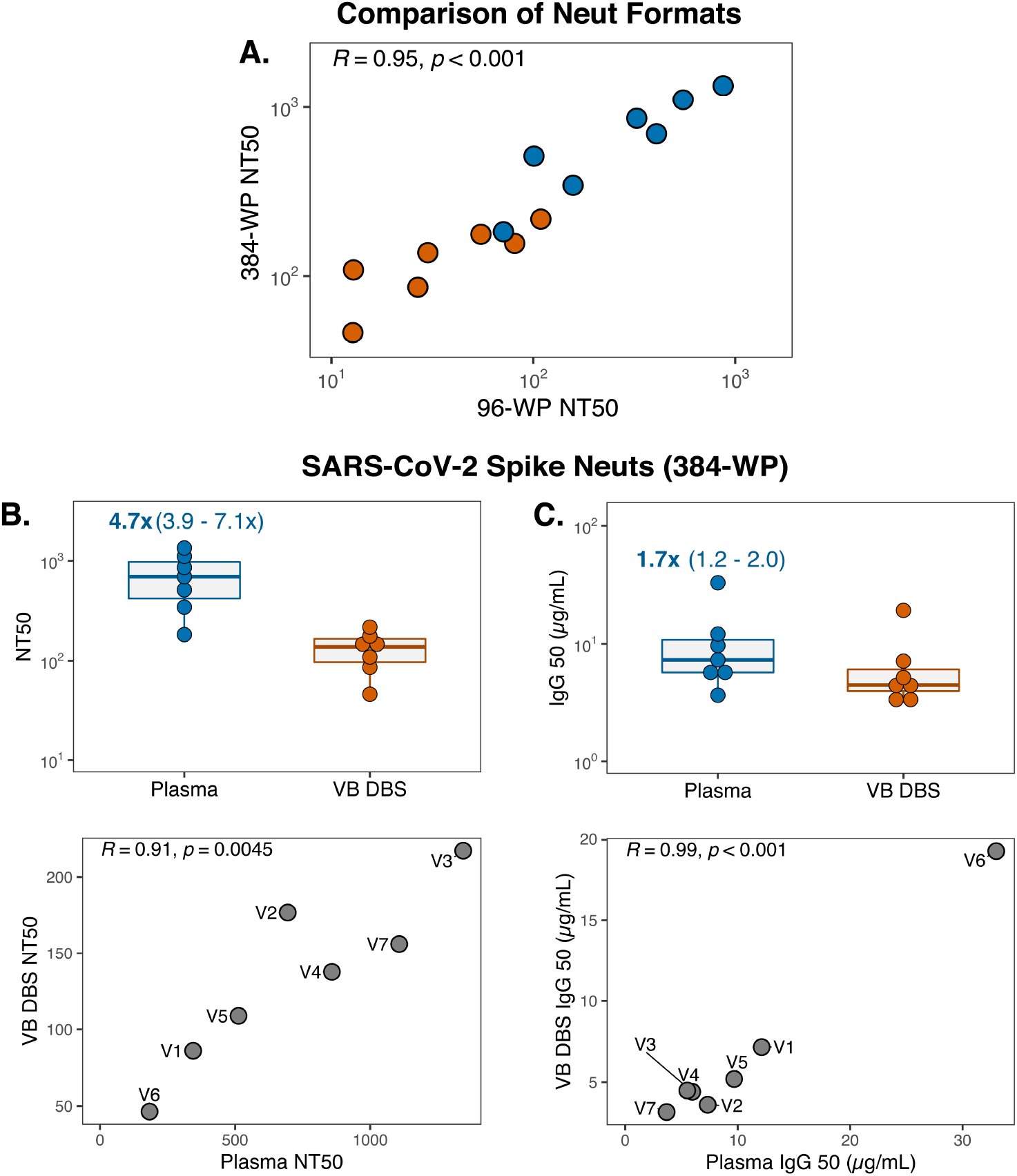
SARS-CoV-2 spike neutralization can be reliably detected in a high-throughput 384 well-plate (WP) format. (**A**) Pearson R correlation of NT_50_ measurements for Group 2 plasma samples (blue) and VB DBS eluates (orange) as determined by 96-WP and 384-WP neutralization assays. (**B**) NT_50_ and (**C**) IgG_50_ results for sample type pairs and Pearson R correlations in the 384-WP format. Data points in the correlation plots are labeled with Group 2 participant IDs (V1-V7).

### Consistent levels of antibody recovery from DBS cards stored at room temperature for six months

One of the main advantages of DBS sampling is that DBS cards do not require cold chain for collection, transportation, or storage. To evaluate the stability of antibodies on DBS cards after prolonged storage without refrigeration or freezing, we measured total IgG and SARS-CoV-2 RBD IgG binding in eluates from Group 1 VB DBS cards after one week, six weeks, and six months of room temperature storage (**Figure 6**). DBS cards were kept in the dark in individual plastic bags with desiccant packets during this time, which is standard practice as humidity and ultraviolet light are known to damage DBS (18). Total IgG recovered from DBS was similar at six months compared to one week post-collection and levels were highly correlated (Pearson R=0.91). Likewise, SARS-CoV-2 RBD binding was consistent over this period (Pearson R=0.97), demonstrating that antibodies are preserved on DBS cards stored in room temperature for at least six months.

**Figure 6.**
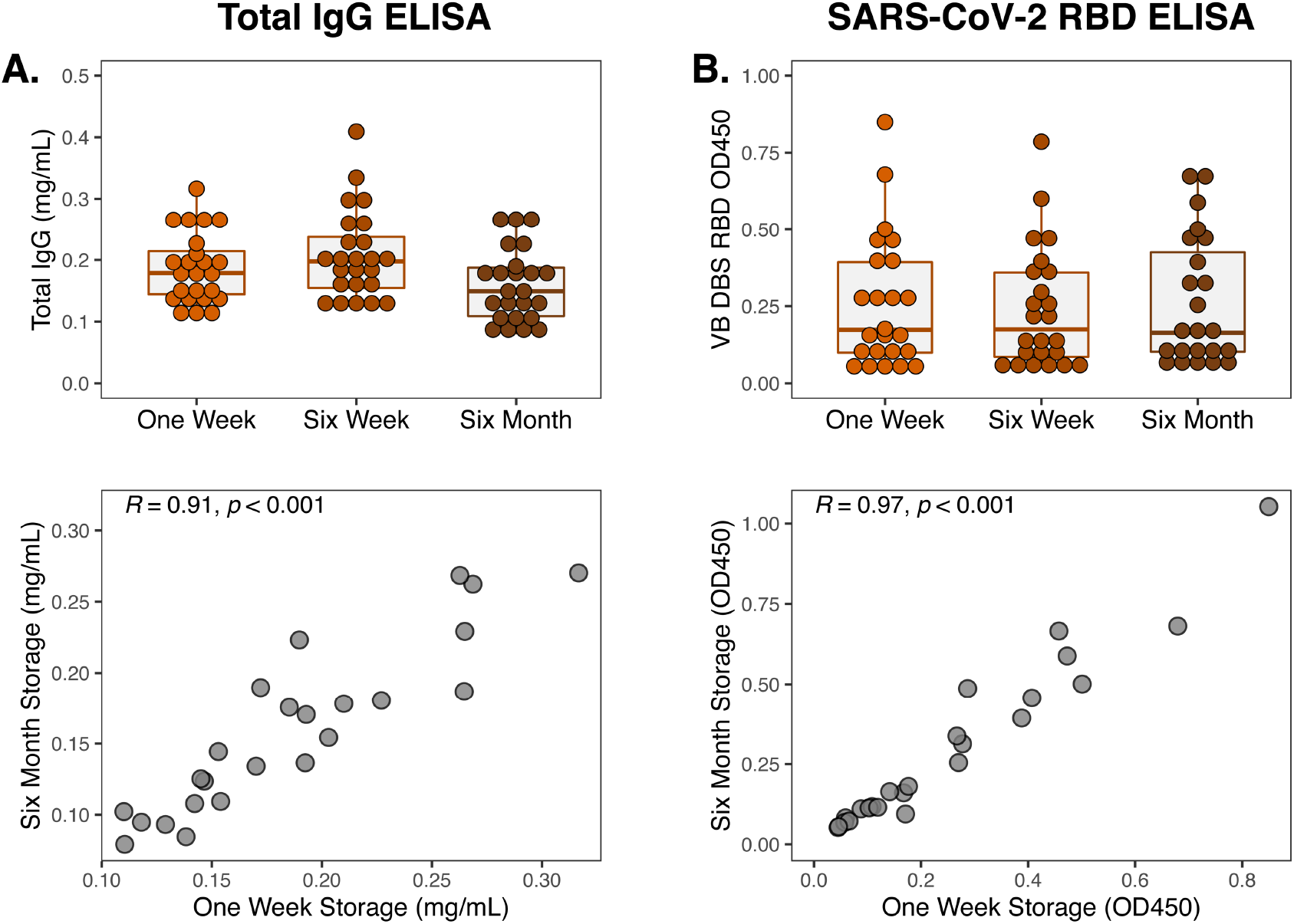
Consistent levels of antibody recovery from DBS cards stored at room temperature for six months. (**A**) Total and (**B**) RBD-specific IgG levels in eluates from Group 1 VB DBS cards (n=24) stored at room temperature for one week, six weeks, and six months. Pearson R correlation between IgG levels measured after storage for one week and for six months are also depicted.

### IgG binding and neutralization activity are highly correlated for self-collected FS DBS and plasma

The DBS cards used so far in this study were prepared with venous blood, similar to prior studies (27, 30, 42-44), and were thus informative intermediates to understand antibody features that can be captured and preserved on filter paper. However, the ideal DBS sample for broad field application would be fingerstick DBS cards (FS DBS) self-collected by individuals at home and mailed to the clinic for storage, processing, and evaluation. To determine whether FS DBS eluates also generate results that reflect plasma, we collected plasma from eight individuals with prior SARS-CoV-2 infection, some of whom were also vaccinated against SARS-CoV-2 (5/8 individuals; Group 3 in **Figure 1**). During their clinic visit for plasma collection, individuals were provided with a kit for FS DBS card preparation (**Figure S2**) and were instructed to fill out the card on the same day, dry it for three hours, and mail it back to the clinic.

Like VB DBS samples, eluates from Group 3 FS DBS cards recapitulated total IgG trends observed in paired plasma (**Figure 7A**; Pearson R=0.92). We then measured RBD IgG binding in these samples, accounting for the 10-fold difference in IgG levels between sample types. As expected, there was no difference in the magnitude of RBD IgG OD measurements (**Figure 7B**). These results were perfectly correlated between sample types (Pearson R=1) and only demonstrated a mean bias of 0.024 OD. In this dataset, we noticed that sample RBD binding was bimodally distributed for both sample types. This difference was driven by vaccination status, as the five individuals with detectable RBD binding all previously received SARS-CoV-2 vaccines whereas participants F1, F4, and F8 had not.

**Figure 7.**
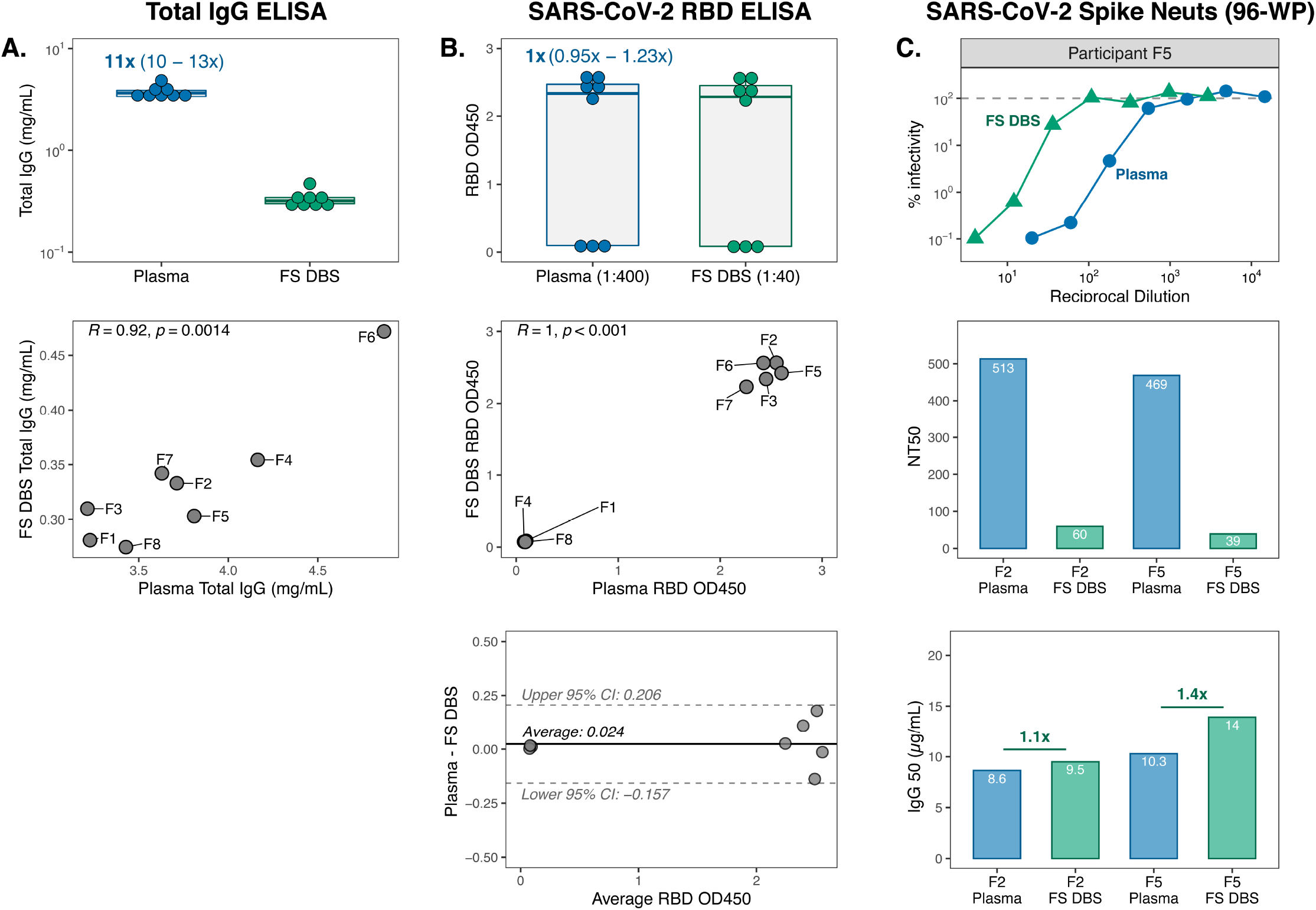
IgG binding and neutralization activity are highly correlated for self-collected FS DBS and plasma. (**A**) Total IgG concentrations and (**B**) RBD binding levels in Group 3 paired sample types (n=8 pairs) with respective Pearson R correlations and Bland-Altman plot analyses. Data points in the correlation plots are labeled with Group 3 participant IDs (F1-F8). (**C**) Neutralization curves against SARS-CoV-2 spike pseudovirus by plasma and FS DBS eluate from participant F5, as well as NT_50_ and IgG_50_ results for paired sample types from participants F2 and F5.

Finally, we assessed spike neutralization activity in two Group 3 participants (F2, F5) who demonstrated high RBD binding and had enough DBS discs remaining. Though this sample size was low, the shape of the neutralization curves was very similar between sample types (**Figure 7C**) and the trend in NT_50_ values was maintained, with participant F2 having slightly higher NT_50_ than F5. IgG_50_ levels were also very similar for these two sample pairs, which suggests that FS DBS, like VB DBS, accurately reflect plasma neutralization activity.

## DISCUSSION

DBS sampling overcomes the logistical barriers of plasma collection because the cards can be self-collected by fingerstick and mailed to the laboratory at ambient temperature, where they are eluted for antibody assays. For SARS-CoV-2 studies, adoption of DBS may reduce frontline worker demand, improve sampling in resource-limited areas, and increase study enrollment. Though prior work with DBS and plasma pairs has supported the suitability of DBS for SARS-CoV-2 antibody binding assays (27-36), important gaps remained that prevented the broad implementation of this sample type. For example, most studies used DBS with venous (27, 30) or fingerstick blood collected at the clinic with the assistance of a phlebotomist (29, 31-33, 35), which does not account for variables introduced with self-sampling or ambient shipping. One study did utilize self-collected fingerstick cards (28), but only evaluated four sample pairs via an agglutination-PCR assay. Finally, all prior reports evaluated antibody binding in COVID-19 convalescent DBS samples. As vaccine distribution continues to expand and evidence grows for neutralization being an important correlate of protection (12, 13, 41), it is critical that DBS eluates also recapitulate SARS-CoV-2 neutralization activity in vaccinated individuals.

In this study, we build on previous findings that supported DBS for antibody binding by collecting and testing paired DBS and plasma from 39 individuals across a range of assays, including epitope profiling and spike neutralization assays. We evaluated not only convalescent samples but also samples from vaccinees and fingerstick DBS cards self-collected at home. The work reported here thus extends the utility of DBS sampling to ultimately support the adoption of DBS for SARS-CoV-2 antibody studies.

Prior studies often elute DBS discs in volumes that prepare samples directly for testing in a single assay format (27, 29, 31, 32, 34, 36). Because we sought to employ DBS for various assays, we first measured total IgG levels in DBS eluate and plasma pairs to estimate the volume of plasma extracted from each disc, to ultimately inform sample dilutions and data normalization approaches. Based on the total IgG results from all 39 sample pairs, we calculated that a median of 6.1 µL plasma was eluted from each 6-mm DBS subpunch. This estimate agrees with a report that compared spotted blood volume to DBS area and determined that 6-mm punches contain 5.8-6.4 µL plasma (27). When we accounted for the dilution factor introduced by eluting the 6.1 µL plasma from each disc into larger volumes, we found that DBS eluates recapitulated not only the trends but also the binding magnitude of paired plasma for binding and neutralization assays. These findings thus demonstrate that plasma and total IgG content on DBS discs can be accounted for during DBS elution, sample dilution, and/or data analysis to ensure assay sensitivity and facilitate direct comparisons between plasma and DBS results.

Our study and prior reports demonstrate strong agreement between paired plasma and DBS for single-epitope SARS-CoV-2 binding assays, such as the commonly used RBD ELISA. However, several epitopes need to be interrogated to elucidate immunodominant proteins or the effects of variant mutations. Phage-display libraries enable multi-epitope investigations by providing high-throughput assessment of binding to linear peptides (45). To determine whether DBS reflect the polyclonal specificities that exist in plasma, we assayed paired samples from 22 convalescent individuals against a recently reported phage library that spans the SARS-CoV-2 proteome (39). DBS eluates identified the same immunodominant proteins as plasma and reflected plasma epitope profiles on both the cohort-level and for individual participants. Individual-level agreement between plasma and DBS was further improved by only considering samples with high reproducibility between replicates, suggesting that replicate experiments may provide the best accuracy.

We also evaluated the performance of DBS in SARS-CoV-2 pseudovirus neutralization assays – the traditional 96-WP format and a new high-throughput 384-WP adaptation. In the 96-WP version, DBS replicated trends in plasma NT_50_ results and matched plasma IgG_50_ levels, demonstrating for the first time that plasma neutralization against SARS-CoV-2 spike is preserved on DBS cards. While these findings are encouraging, the 96-WP neutralization assay is not ideal for testing DBS eluates or precious plasma specimens due to large sample volume requirements. We therefore established a 384-WP format for the assessment of both plasma and DBS eluates that uses a third of the sample volume and can assay five times the number of samples per plate. NT_50_ results strongly correlated between assay formats and the agreement between DBS and plasma was maintained. The 384-WP SARS-CoV-2 spike neutralization assay is thus a sample-sparing, high-throughput approach for the evaluation of plasma and DBS eluates.

Our study ultimately provides evidence that SARS-CoV-2 antibody binding and neutralization are detectable in eluates from DBS cards, including those prepared at home by fingerstick and mailed to the clinic, at levels that reflect paired plasma. This work supports the implementation of DBS sampling for SARS-CoV-2 antibody investigations as a more practical alternative to plasma and sera collection, particularly in resource-limited settings.

## METHODS

### Study participants

Paired DBS and plasma specimens were collected from COVID-19 convalescent and SARS-CoV-2 vaccinated individuals enrolled as part of the Hospitalized or Ambulatory Adults with Respiratory Viral Infections (HAARVI) research study in Seattle, Washington. All participants completed informed consent as approved by the University of Washington Institutional Review Board (protocol #STUDY00000959). Paired samples from 39 HAARVI participants were collected between September 2020 and February 2021 and were categorized into three sample groups for this study depending on the individual’s SARS-CoV-2 infection and vaccination history and the specimen types collected (**Figure 1**). Group 1 consisted of 24 convalescent individuals with paired plasma and VB DBS cards. Group 2 included seven vaccinated participants with paired plasma and VB DBS cards. Finally, Group 3 was comprised of eight convalescent individuals, five of whom were also vaccinated, with paired plasma and FS DBS cards. Convalescent individuals refer to those with previous PCR-confirmed, symptomatic SARS-CoV-2 infection that did not require hospitalization. Vaccinated participants received two doses of the Moderna mRNA-1273 or Pfizer/BioNTech BNT162b2 SARS-CoV-2 vaccine prior to sample collection.

### Plasma and VB DBS sample collection

Venous blood was collected from each participant into acid citrate dextrose (ACD) tubes. For individuals in Groups 1 and 2, VB DBS cards were prepared immediately after venipuncture by inverting blood tubes, pipetting 80 µL onto each circle of a Whatman 903 card (Sigma #WHA10534612), and drying cards overnight at room temperature (RT). Dried VB DBS cards were stored in a drawer at RT in individual plastic bags containing a desiccant packet (Grainger #8ZF81). All plasma specimens were heat-inactivated at 56ºC for one hour prior to short-term storage at 4ºC or long-term storage at -80ºC.

### At-home FS DBS collection kits

Group 3 participants were provided with FS DBS collection kits and instructions at the time of an in-clinic venous blood draw and were advised to perform FS DBS sampling on the same day. Kits included the following: an instruction pamphlet (**Figure S2**), one Whatman 903 card in a plastic bag with a desiccant packet, two lancets, one alcohol prep pad, one 95kPA specimen transport bag, one cardboard box, and one prepaid mailer bag. FS DBS cards were mailed to the laboratory at ambient temperature and were stored at RT in the same conditions as VB DBS cards.

### DBS elution

Six-mm discs were extracted from saturated portions of VB or FS DBS cards using a biopsy punch (4MD Medical #MLTX33-36). For FS DBS cards with variable spot sizes, discs were extracted from spots that were at least 6-mm in diameter to ensure a fully saturated sample. Forceps were used to transfer discs into a sterile two-mL Eppendorf tube. To prevent cross-contamination, biopsy punches were replaced between cards and forceps were cleaned with 70% ethanol. PBS-T (1X PBS, 0.1% Tween-20) was added to each tube at a ratio of one disc to 100 µL PBS-T unless otherwise noted. Samples were eluted overnight at 4ºC on a plate shaker with gentle agitation (70 RPM). In the morning, tubes were centrifuged at 10,000 x g for 5 minutes at RT to pellet discs and debris. Supernatants were extracted and stored at 4ºC.

### Quantification of total IgG concentrations

ELISAs were performed to determine total IgG levels in paired plasma and DBS eluates. Immulon 2HB 96-well plates (ThermoFisher #3455) were coated with 50 µL of 25 µg/mL goat anti-human IgGs polyvalent (Sigma #I1761) in 0.1M sodium bicarbonate at 4ºC overnight. Wells were washed three times with 300 µL PBS-T using a Tecan Plate Washer and blocked with 50 µL blocking buffer (10% w/v non-fat milk and 0.05% Tween-20 in 1X PBS) for one hour at RT. Plasma and DBS eluates were briefly centrifuged at 10,000 x g for 5 minutes at RT to pellet debris. Sample dilutions were then prepared in blocking buffer (1:10,000 for plasma, 1:5,000 for VB DBS, 1:10,000 for FS DBS). An IgG antibody of known concentration was diluted to 3 µg/mL and was titrated three-fold across 10 wells to serve as the assay standard. Blocking buffer was washed from assay plates and 50 µL of samples and standards were added to plates in duplicate. After one hour at RT, wells were washed three times. Secondary antibody was prepared by diluting goat anti-human IgG-horseradish peroxidase (Sigma #A0170) to 1:2,500 in blocking buffer (2.24 µg/mL). 100 µL was added per well and plates were incubated at RT for one hour. After incubation with secondary antibody and three washes, plates were developed with 50 µL of TMB substrate (ThermoFisher #34029) and were quenched with equal volumes of 1N sulfuric acid after ten minutes. Absorbance was immediately read at 450 nm on a BioTek Epoch plate reader. Duplicate OD_450_ measurements were averaged and IgG concentrations in plasma and DBS eluates were interpolated from the standard curve using the five-parameter logistic equation function in GraphPad Prism 8. Agreement between paired sample IgG concentrations was evaluated by calculating the Pearson correlation coefficient between sample types in RStudio.

### Detection of SARS-CoV-2 RBD IgG

A previously described IgG ELISA against SARS-CoV-2 RBD that gained FDA emergency use authorization was adapted to measure RBD binding activity in paired plasma and DBS eluates (38). Immulon 2HB 96-well plates (ThermoFisher #3455) were coated with 50 µL/well of 2 µg/mL of SARS-CoV-2 RBD protein diluted in 1X PBS. The RBD protein used as coat was a gift from Dr. Roland Strong’s lab and was produced as previously described (46). After an overnight incubation at 4ºC, plates were washed three times with 300 µL PBS-T using a Tecan Plate Washer and blocked with 200 µL/well of 3% w/v non-fat milk in PBS-T for one hour at RT. Plasma and DBS eluates were centrifuged at 10,000 x g for 5 minutes at RT to pellet debris prior to being diluted in dilution buffer (1% w/v non-fat milk in PBS-T). The following point dilutions were assessed: 1:400 for participant plasma and negative control normal human serum (Gemini Biosciences #100-110, lot H87WOOK), 1:20 for VB DBS eluates, and 1:40 for FS DBS eluates. RBD-specific CR3022 IgG (BEI Resources #NR-52392) was used as a positive control at 1 µg/mL. After the blocking buffer was removed, 100 µL of sample dilutions were added in duplicate and assay plates were incubated for 2 hours at RT. Wells were washed three times with PBS-T and 50 µL of goat anti-human IgG-horseradish peroxidase (Sigma #A0170) diluted 1:3000 in dilution buffer (1.87 µg/mL) was added per well. After 1 hour at RT, plates were washed three times with PBS-T. 50 µL of TMB substrate (ThermoFisher #34029) per well was used to develop assay plates, and after 5 minutes 50 µL of 1N sulfuric acid was used to stop the reaction. Absorbance was immediately read at 450 nm on a BioTek Epoch plate reader. Agreement between paired sample RBD OD450 measurements was evaluated by calculating the Pearson correlation coefficient between sample types in RStudio.

### Linear epitope mapping via phage display and immunoprecipitation

Linear CoV epitope profiling of paired plasma and VB DBS eluates was performed directly following a phage-display approach previously described in detail by our group (39). Specifically, we employed the same pan CoV phage library design and construction, IgA and IgG immunoprecipitation, Illumina library preparation, and sequence alignment techniques as the previous report. This pan CoV phage library displays peptides 39 amino acids in length that tile across 17 CoV protein coding sequences, including the entire SARS-CoV-2-Wuhan-1 proteome (GenBank MN908947). For epitope mapping experiments, plasma and VB DBS eluates are added to wells containing the phage library such that each well contains 10 µg of total IgG, as estimated by ELISA results. After rotating for 20 hours at 4ºC, phage-antibody complexes were pulled down by immunoprecipitation using a 1:1 ratio of protein A and G Dynabeads (Invitrogen #10002D and #10004D) and were lysed at 95ºC for ten minutes.

Sample-selected library phage DNA was prepared for sequencing on an Illumina MiSeq with 126 bp single-end reads using the methods and primers previously described (47). Sequences were aligned to the pan CoV reference library and counts per million (CPM) values were calculated from raw read counts to control for read depth differences between samples. Due to the focused nature of this investigation on whether DBS agrees with plasma for SARS-CoV-2 antibody responses, downstream analyses specifically evaluated the 480 SARS-CoV-2 peptides included in the library. The following analyses were performed in R to compare plasma and VB DBS linear epitope profiling results: (1) Pearson correlation coefficients were determined for CPMs between within-assay technical replicates to inform data quality. (2) CPMs for within-assay replicates were then averaged and participant-level agreement between sample types was assessed by comparing CPMs for VB DBS and plasma pairs via Pearson correlation. (3) Cohort-level agreement between sample types was evaluated by calculating the average CPM for each peptide across all plasma or VB DBS samples and determining the Pearson correlation coefficient between sample type averages.

### SARS-CoV-2 spike pseudovirus production

Pseudovirus expressing SARS-CoV-2 spike protein was produced and titered as previously described (40). HEK293T cells were added to 6-well plates at 5 x 10^5^ cells per well in DMEM supplemented with 10% fetal bovine serum, 2 mM L-glutamine, and penicillin/streptomycin/fungizone. After 16-25 hours, cells were transfected using FuGENE-6 (Promega #E2692) with the Luciferase_IRES_ZsGreen backbone, Gag/Pol, Rev, and Tat lentiviral helper plasmids, and a plasmid containing the codon-optimized spike sequence from the Wuhan-Hu-1 strain which contained a 21 amino acid deletion at the cytoplasmic tail (also known as HDM-SARS2-Spike-delta21). After 24 hours, media was replaced with fresh supplemented DMEM. Between 50-60 hours post-transfection, viral supernatants were collected, filtered through 0.22 µm Steriflip filters, concentrated using 100KDa Amicon filters (EMD Millipore # UFC910024), and stored at -80ºC. Pseudovirus was titered by seeding 1.25 x 10^4^ HEK293T-ACE2 cells in 96-well black-walled plates and, after 16-24 hours, adding 100 µL of diluted viral supernatant per well in duplicate. Viral supernatants were initially diluted 1:10 in supplemented DMEM and were then diluted two-fold over seven wells. VSV-G and no Viral Entry Protein (VEP) positive and negative controls, respectively, were included on each plate and treated the same as spike pseudovirus except that VSV-G titration started at a 1:10 dilution. After 60 hours, 100 µL per well was removed and 30 µL of Bright-Glo (Promega #E2620) was added. Relative luciferase units (RLU) were measured on a LUMIstar Omega plate reader (BMG Labtech).

### SARS-CoV-2 spike neutralization assays (96-WP and 384-WP)

DBS samples were eluted for neutralization assays with 50 µL of serum-free supplemented DMEM to ensure their compatibility with cell culture. SARS-CoV-2 spike neutralization assays in the traditional 96-WP format were carried out according to a previous report (40). HEK293T-ACE2 cells were brought to 2.5 x 10^5^ cells/mL and 50 µL was seeded in 96-well black walled plates. Assay plates also contained four wells seeded with HEK293T cells without ACE2 and four wells without any cells as negative controls. After 12-16 hours, 60 µL of plasma and DBS dilutions were prepared in a separate round-bottomed 96-WP plate with serum-free supplemented DMEM, due to the fact that DBS were eluted in this media. Samples were initially diluted 1:20 and 1:4 for plasma and DBS, respectively, and were then diluted three-fold over seven wells. Duplicate dilution wells were prepared for each sample titration. Spike pseudovirus was then diluted to 3.3-5 x 10^5^ RLU per mL in supplemented DMEM and 60 µL was mixed with the prepared plasma and DBS titrations. After 1 hour at 37ºC, 100 µL of the virus/sample wells was transferred to the cell plate. To read the plates, 100 µL of media was removed from each well approximately 52-58 hours post-infection and 30 µL of Bright-Glo was added. After two minutes, RLU was measured on a LUMIstar Omega plate reader. Technical replicate RLUs were averaged and percent infectivity was calculated by dividing sample RLU by the corresponding row’s positive control RLU, which was from virus plus cells wells that did not have sample added. Neutralization titers (NT_50_) were determined using GraphPad Prism 8’s inhibitor versus response curve with top and bottom parameters constrained to 1 and 0, respectively. Finally, IgG_50_ values were calculated by dividing each sample’s total IgG concentration by its NT_50_. Agreement between paired sample measurements for NT_50_ and IgG_50_ values was evaluated by calculating the Pearson correlation coefficient between sample types in RStudio.

Spike neutralization assays were adapted to 384-WPs using the same incubation times, sample dilutions, and reagents as described for the 96-WP assay, but with working volumes reduced to approximately 30%. Briefly, black-walled 384-WPs were seeded with 15 µL of HEK293T-ACE2 cells at a concentration of 2.5 x 10^5^ cells/mL. For negative controls, eight wells were seeded with HEK293T cells that do not express ACE2 and an additional eight wells contained supplemented DMEM only. After 12-16 hours, sample dilutions were prepared in round-bottomed 96-WPs with a total volume of 36 µL per well. Spike pseudovirus was diluted to 3.3-5 x 10^5^ RLU per mL and 36 µL was mixed into sample dilution wells. After 1 hour at 37ºC, 30 µL from each virus/sample well was transferred to two wells on the 384-well assay plate. Roughly 52-58 hours post-infection, plates were read by removing 30 µL of media per well, adding 9 µL of BrightGlo per well, and measuring RLU on a LUMIstar Omega plate reader after 2 minutes.

### Quantification and statistical analysis

Illumina MiSeq Reporter software was used by the by the Fred Hutch Genomics Core to demultiplex peptide epitope-mapping sequencing data and generate fastq files. We then used a Nextflow data processing pipeline to align the demultiplexed sample reads to the reference peptide library in parallel, allowing for up to two mismatches. This pipeline builds a Bowtie index from the peptide metadata by converting the metadata to fasta format and feeding it into the bowtie-build command. The low-quality end of the reads is trimmed to 93bp to match the reference lengths before performing end-to-end alignment and allowing for 0 mismatches. For each sample, we quantified the abundance of each peptide by using samtools-idxstats to count the number of reads mapped to each specific peptide in the reference library. The peptide counts were merged into a count matrix organized by unique identifiers for each peptide and sample. The metadata tables were tied with the count matrix into an xarray dataset using shared coordinate dimensions of the unique sample and peptide identifiers. We used this dataset organization as the starting point for downstream epitope-profiling analyses.

Additional statistical analyses were performed using RStudio or GraphPad Prism 8. Figure 1 was created in BioRender.com. The remaining figures were generated using RStudio. Pearson R correlations were used to compare trends between DBS eluates and paired plasma for all assays. All data represent the mean of two technical replicates. Statistical significance was defined as a p-value less than 0.05. Additional details can be found in the figure legends and corresponding results sections.

## Data Availability

The Nextflow pipeline, used to align epitope-mapping sample reads to the reference library, is available at https://github.com/matsengrp/phip-flow. The custom RStudio code generated and used in this study to perform statistical analyses and visualize data is available upon request. Any additional information required to reanalyze the data reported in this paper is also available upon request.

## ACKNOWLEDGEMENTS

We thank the dedicated HAARVI staff and all participants who contributed samples. We also thank members of the Overbaugh, Matsen, and Chu labs for helpful discussion and feedback. This work was supported by the following NIH grants: R01 AI138709 awarded to J.O., T32 GM007270 awarded to H.L.I., BMGF Acceleration of COVID-19 Research to H.Y.C., and R01 AI146028 awarded to F.A.M.. J.O. also received support as the Endowed Chair for Graduate Education (Fred Hutch). The research of F.A.M. was supported in part by a Faculty Scholar grant from the Howard Hughes Medical Institute and the Simons Foundation. Scientific Computing Infrastructure at Fred Hutch funded by ORIP grant S10OD028685.

## SUPPLEMENTAL INFORMATION

**Figure S1.**
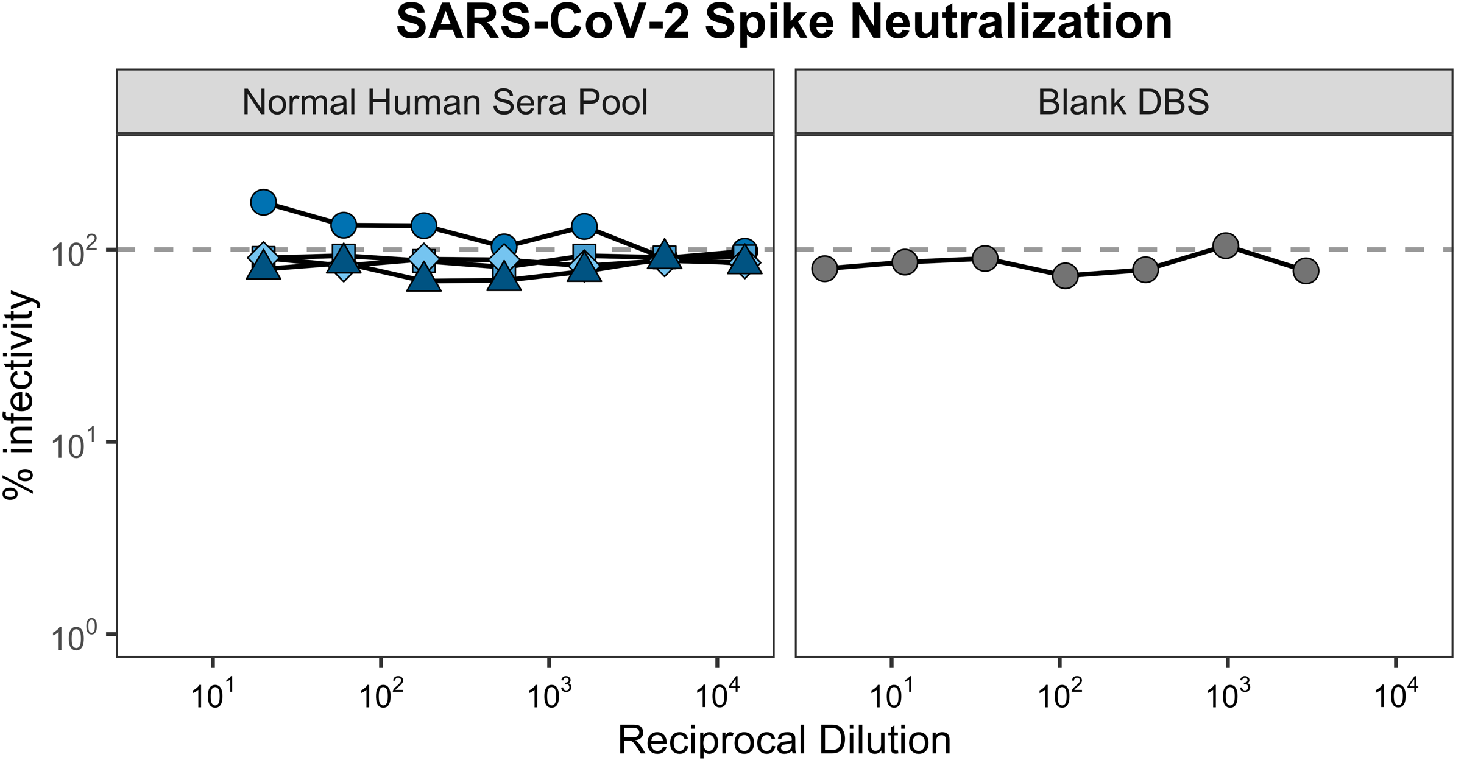
Undetectable levels of spike neutralization in negative control DBS and plasma. A pre-pandemic normal human sera pool (n=4 assay days) and the eluate from a blank DBS card were tested for SARS-CoV-2 spike neutralization activity at the same dilutions as participant samples, according to sample type.

**Figure S2.**
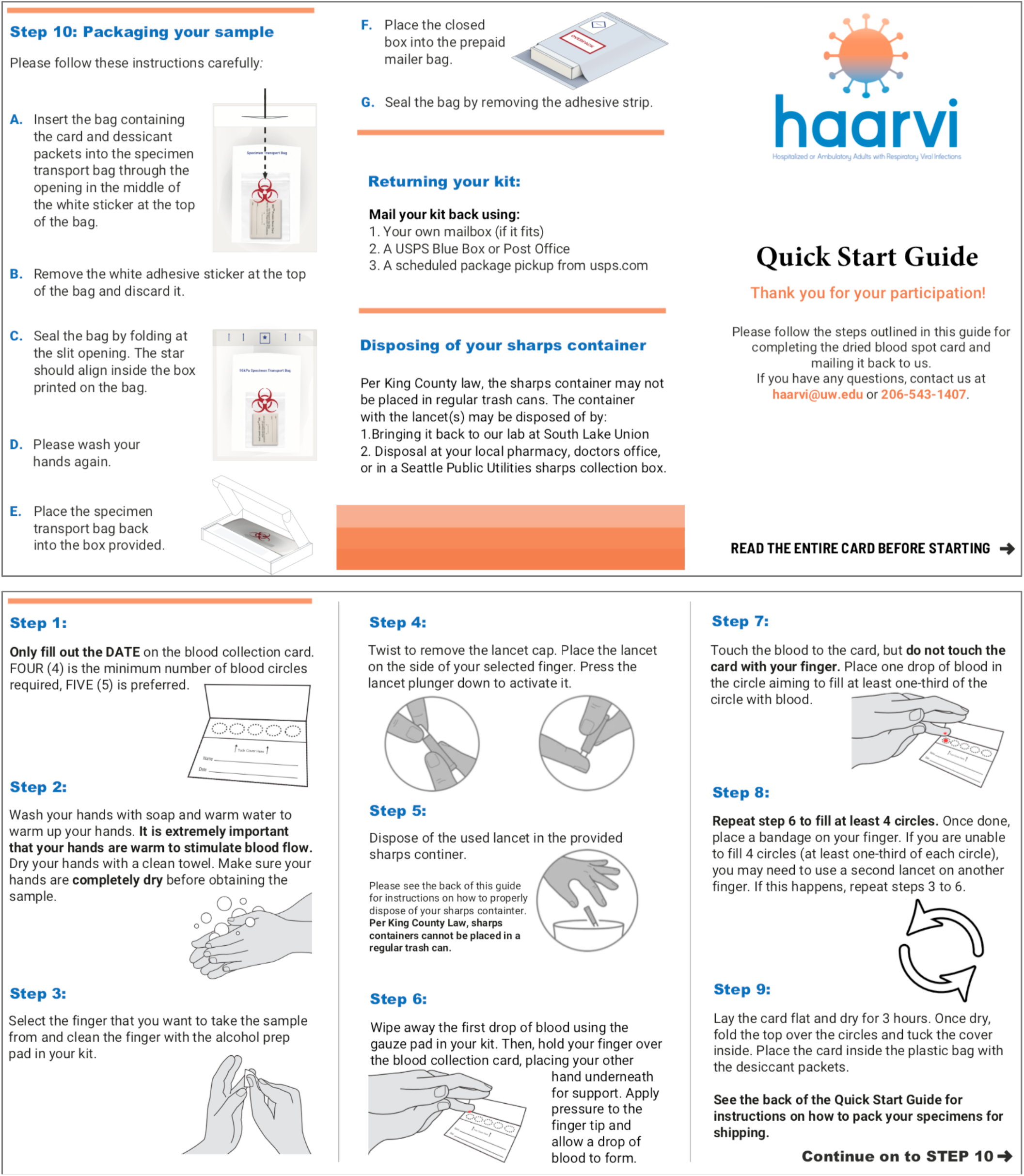
Instruction pamphlet for self-collection of fingerstick DBS cards. Study participants were provided with the depicted instructions and materials during a clinic visit where a blood sample was collected.

## REFERENCES

1. Zhang Z, Bi Q, Fang S, Wei L, Wang X, He J, Wu Y, Liu X, Gao W, Zhang R, Gong W, Su Q, Azman AS, Lessler J, Zou X (2021). Insight into the practical performance of RT-PCR testing for SARS-CoV-2 using serological data: a cohort study. Lancet Microbe 2(2): e79-e87. PMID: 33495759. PMCID: PMC7816573.

2. Okba NMA, Muller MA, Li W, Wang C, GeurtsvanKessel CH, Corman VM, Lamers MM, Sikkema RS, de Bruin E, Chandler FD, Yazdanpanah Y, Le Hingrat Q, Descamps D, Houhou-Fidouh N, Reusken C, Bosch BJ, Drosten C, Koopmans MPG, Haagmans BL (2020). Severe Acute Respiratory Syndrome Coronavirus 2-Specific Antibody Responses in Coronavirus Disease Patients. Emerg Infect Dis 26(7): 1478-88. PMID: 32267220. PMCID: PMC7323511.

3. Seow J, Graham C, Merrick B, Acors S, Pickering S, Steel KJA, Hemmings O, O’Byrne A, Kouphou N, Galao RP, Betancor G, Wilson HD, Signell AW, Winstone H, Kerridge C, Huettner I, Jimenez-Guardeno JM, Lista MJ, Temperton N, Snell LB, Bisnauthsing K, Moore A, Green A, Martinez L, Stokes B, Honey J, Izquierdo-Barras A, Arbane G, Patel A, Tan MKI, O’Connell L, O’Hara G, MacMahon E, Douthwaite S, Nebbia G, Batra R, Martinez-Nunez R, Shankar-Hari M, Edgeworth JD, Neil SJD, Malim MH, Doores KJ (2020). Longitudinal observation and decline of neutralizing antibody responses in the three months following SARS-CoV-2 infection in humans. Nat Microbiol 5(12): 1598-607. PMID: 33106674. PMCID: PMC7610833.

4. Lumley SF, O’Donnell D, Stoesser NE, Matthews PC, Howarth A, Hatch SB, Marsden BD, Cox S, James T, Warren F, Peck LJ, Ritter TG, de Toledo Z, Warren L, Axten D, Cornall RJ, Jones EY, Stuart DI, Screaton G, Ebner D, Hoosdally S, Chand M, Crook DW, O’Donnell AM, Conlon CP, Pouwels KB, Walker AS, Peto TEA, Hopkins S, Walker TM, Jeffery K, Eyre DW, Oxford University Hospitals Staff Testing G (2021). Antibody Status and Incidence of SARS-CoV-2 Infection in Health Care Workers. N Engl J Med 384(6): 533-40. PMID: 33369366. PMCID: PMC7781098.

5. Ebinger JE, Fert-Bober J, Printsev I, Wu M, Sun N, Prostko JC, Frias EC, Stewart JL, Van Eyk JE, Braun JG, Cheng S, Sobhani K (2021). Antibody responses to the BNT162b2 mRNA vaccine in individuals previously infected with SARS-CoV-2. Nat Med 27(6): 981-4. PMID: 33795870. PMCID: PMC8205849.

6. Krammer F, Srivastava K, Alshammary H, Amoako AA, Awawda MH, Beach KF, Bermudez-Gonzalez MC, Bielak DA, Carreno JM, Chernet RL, Eaker LQ, Ferreri ED, Floda DL, Gleason CR, Hamburger JZ, Jiang K, Kleiner G, Jurczyszak D, Matthews JC, Mendez WA, Nabeel I, Mulder LCF, Raskin AJ, Russo KT, Salimbangon AT, Saksena M, Shin AS, Singh G, Sominsky LA, Stadlbauer D, Wajnberg A, Simon V (2021). Antibody Responses in Seropositive Persons after a Single Dose of SARS-CoV-2 mRNA Vaccine. N Engl J Med 384(14): 1372-4. PMID: 33691060. PMCID: PMC8008743.

7. Widge AT, Rouphael NG, Jackson LA, Anderson EJ, Roberts PC, Makhene M, Chappell JD, Denison MR, Stevens LJ, Pruijssers AJ, McDermott AB, Flach B, Lin BC, Doria-Rose NA, O’Dell S, Schmidt SD, Neuzil KM, Bennett H, Leav B, Makowski M, Albert J, Cross K, Edara VV, Floyd K, Suthar MS, Buchanan W, Luke CJ, Ledgerwood JE, Mascola JR, Graham BS, Beigel JH, m RNASG (2021). Durability of Responses after SARS-CoV-2 mRNA-1273 Vaccination. N Engl J Med 384(1): 80-2. PMID: 33270381. PMCID: PMC7727324.

8. Baden LR, El Sahly HM, Essink B, Kotloff K, Frey S, Novak R, Diemert D, Spector SA, Rouphael N, Creech CB, McGettigan J, Khetan S, Segall N, Solis J, Brosz A, Fierro C, Schwartz H, Neuzil K, Corey L, Gilbert P, Janes H, Follmann D, Marovich M, Mascola J, Polakowski L, Ledgerwood J, Graham BS, Bennett H, Pajon R, Knightly C, Leav B, Deng W, Zhou H, Han S, Ivarsson M, Miller J, Zaks T, Group CS (2021). Efficacy and Safety of the mRNA-1273 SARS-CoV-2 Vaccine. N Engl J Med 384(5): 403-16. PMID: 33378609. PMCID: PMC7787219.

9. Polack FP, Thomas SJ, Kitchin N, Absalon J, Gurtman A, Lockhart S, Perez JL, Perez Marc G, Moreira ED, Zerbini C, Bailey R, Swanson KA, Roychoudhury S, Koury K, Li P, Kalina WV, Cooper D, Frenck RW, Jr., Hammitt LL, Tureci O, Nell H, Schaefer A, Unal S, Tresnan DB, Mather S, Dormitzer PR, Sahin U, Jansen KU, Gruber WC, Group CCT (2020). Safety and Efficacy of the BNT162b2 mRNA Covid-19 Vaccine. N Engl J Med 383(27): 2603-15. PMID: 33301246. PMCID: PMC7745181.

10. Sadoff J, Gray G, Vandebosch A, Cardenas V, Shukarev G, Grinsztejn B, Goepfert PA, Truyers C, Fennema H, Spiessens B, Offergeld K, Scheper G, Taylor KL, Robb ML, Treanor J, Barouch DH, Stoddard J, Ryser MF, Marovich MA, Neuzil KM, Corey L, Cauwenberghs N, Tanner T, Hardt K, Ruiz-Guinazu J, Le Gars M, Schuitemaker H, Van Hoof J, Struyf F, Douoguih M, Group ES (2021). Safety and Efficacy of Single-Dose Ad26.COV2.S Vaccine against Covid-19. N Engl J Med 384(23): 2187-201. PMID: 33882225. PMCID: PMC8220996.

11. Voysey M, Clemens SAC, Madhi SA, Weckx LY, Folegatti PM, Aley PK, Angus B, Baillie VL, Barnabas SL, Bhorat QE, Bibi S, Briner C, Cicconi P, Collins AM, Colin-Jones R, Cutland CL, Darton TC, Dheda K, Duncan CJA, Emary KRW, Ewer KJ, Fairlie L, Faust SN, Feng S, Ferreira DM, Finn A, Goodman AL, Green CM, Green CA, Heath PT, Hill C, Hill H, Hirsch I, Hodgson SHC, Izu A, Jackson S, Jenkin D, Joe CCD, Kerridge S, Koen A, Kwatra G, Lazarus R, Lawrie AM, Lelliott A, Libri V, Lillie PJ, Mallory R, Mendes AVA, Milan EP, Minassian AM, McGregor A, Morrison H, Mujadidi YF, Nana A, O’Reilly PJ, Padayachee SD, Pittella A, Plested E, Pollock KM, Ramasamy MN, Rhead S, Schwarzbold AV, Singh N, Smith A, Song R, Snape MD, Sprinz E, Sutherland RK, Tarrant R, Thomson EC, Torok ME, Toshner M, Turner DPJ, Vekemans J, Villafana TL, Watson MEE, Williams CJ, Douglas AD, Hill AVS, Lambe T, Gilbert SC, Pollard AJ, Oxford CVTG1 (2021). Safety and efficacy of the ChAdOx1 nCoV-19 vaccine (AZD1222) against SARS-CoV-2: an interim analysis of four randomised controlled trials in Brazil, South Africa, and the UK. Lancet 397(10269): 99-111. PMID: 33306989. PMCID: PMC7723445.

12. Khoury DS, Cromer D, Reynaldi A, Schlub TE, Wheatley AK, Juno JA, Subbarao K, Kent SJ, Triccas JA, Davenport MP (2021). Neutralizing antibody levels are highly predictive of immune protection from symptomatic SARS-CoV-2 infection. Nat Med. PMID: 34002089.

13. Krammer F (2021). Correlates of protection from SARS-CoV-2 infection. Lancet 397(10283): 1421-3. PMID: 33844964. PMCID: PMC8040540.

14. Krammer F (2020). SARS-CoV-2 vaccines in development. Nature 586(7830): 516-27. PMID: 32967006.

15. Liu X, Shaw R, Stuart ASV, Greenland M, Dinesh T, Provstgaard-Morys S, Clutterbuck E, Ramasamy MN, Aley PK, Farooq Mujadidi Y, Long F, Plested E, Robinson H, Singh N, Walker LL, White R, Andrews N, Cameron JC, Collins AM, Ferreira DM, Hill HC, Green CA, Hallis B, Heath PT, Faust SN, Finn A, Lambe T, Lazarus R, Libri V, Ramsay ME, Read RC, Turner DPJ, Turner PJ, Nguyen-Van-Tam JS, Snape MD, Group C-CS (2021). Safety and Immunogenicity Report from the Com-COV Study – a Single-Blind Randomised Non-Inferiority Trial Comparing Heterologous And Homologous Prime-Boost Schedules with An Adenoviral Vectored and mRNA COVID-19 Vaccine. Preprints with the Lancet.

16. Wu K, Choi A, Koch M, Elbashir S, Ma L, Lee D, Woods A, Henry C, Palandjian C, Hill A, Quinones J, Nunna N, O’Connell S, McDermott AB, Falcone S, Narayanan E, Colpitts T, Bennett H, Corbett KS, Seder R, Graham BS, Stewart-Jones GB, Carfi A, Edwards DK (2021). Variant SARS-CoV-2 mRNA vaccines confer broad neutralization as primary or booster series in mice. bioRxiv. PMID: 33880468. PMCID: PMC8057233.

17. Lim MD (2018). Dried Blood Spots for Global Health Diagnostics and Surveillance: Opportunities and Challenges. Am J Trop Med Hyg 99(2): 256-65. PMID: 29968557. PMCID: PMC6090344.

18. World Health O. WHO manual for HIV drug resistance testing using dried blood spot specimens, third edition. World Health Organization, 2020.

19. IATA (2021). International Air Transport Association: Dangerous Goods Regulations. 62.

20. Gruner N, Stambouli O, Ross RS (2015). Dried blood spots--preparing and processing for use in immunoassays and in molecular techniques. J Vis Exp(97). PMID: 25867233. PMCID: PMC4397000.

21. Guthrie R, Susi A (1963). A Simple Phenylalanine Method for Detecting Phenylketonuria in Large Populations of Newborn Infants. Pediatrics 32: 338-43. PMID: 14063511.

22. Li W, Lee MS. Dried blood spots: Applications and techniques: John Wiley & Sons; 2014.

23. Freeman JD, Rosman LM, Ratcliff JD, Strickland PT, Graham DR, Silbergeld EK (2018). State of the Science in Dried Blood Spots. Clin Chem 64(4): 656-79. PMID: 29187355.

24. Smit PW, Elliott I, Peeling RW, Mabey D, Newton PN (2014). An overview of the clinical use of filter paper in the diagnosis of tropical diseases. Am J Trop Med Hyg 90(2): 195-210. PMID: 24366501. PMCID: PMC3919219.

25. Snijdewind IJ, van Kampen JJ, Fraaij PL, van der Ende ME, Osterhaus AD, Gruters RA (2012). Current and future applications of dried blood spots in viral disease management. Antiviral Res 93(3): 309-21. PMID: 22244848.

26. Bertagnolio S, Parkin NT, Jordan M, Brooks J, Garcia-Lerma JG (2010). Dried blood spots for HIV-1 drug resistance and viral load testing: A review of current knowledge and WHO efforts for global HIV drug resistance surveillance. AIDS Rev 12(4): 195-208. PMID: 21179184.

27. Moat SJ, Zelek WM, Carne E, Ponsford MJ, Bramhall K, Jones S, El-Shanawany T, Wise MP, Thomas A, George C, Fegan C, Steven R, Webb R, Weeks I, Morgan BP, Jolles S (2021). Development of a high-throughput SARS-CoV-2 antibody testing pathway using dried blood spot specimens. Ann Clin Biochem 58(2): 123-31. PMID: 33269949. PMCID: PMC7844389.

28. Karp DG, Danh K, Espinoza NF, Seftel D, Robinson PV, Tsai CT (2020). A serological assay to detect SARS-CoV-2 antibodies in at-home collected finger-prick dried blood spots. Sci Rep 10(1): 20188. PMID: 33214612. PMCID: PMC7678827.

29. Weisser H, Steinhagen K, Hocker R, Borchardt-Loholter V, Anvari O, Kern PM (2021). Evaluation of dried blood spots as alternative sampling material for serological detection of anti-SARS-CoV-2 antibodies using established ELISAs. Clin Chem Lab Med 59(5): 979-85. PMID: 33554537.

30. Toh ZQ, Higgins RA, Anderson J, Mazarakis N, Do LAH, Rautenbacher K, Ramos P, Dohle K, Tosif S, Crawford N, Mulholland K, Licciardi PV (2021). The use of dried blood spots for the serological evaluation of SARS-CoV-2 antibodies. J Public Health (Oxf). PMID: 33611565. PMCID: PMC7928805.

31. Mulchandani R, Brown B, Brooks T, Semper A, Machin N, Linley E, Borrow R, Wyllie D, Investigators E-HS (2021). Use of dried blood spot samples for SARS-CoV-2 antibody detection using the Roche Elecsys (R) high throughput immunoassay. J Clin Virol 136: 104739. PMID: 33588354. PMCID: PMC7817498.

32. Zava TT, Zava DT (2021). Validation of dried blood spot sample modifications to two commercially available COVID-19 IgG antibody immunoassays. Bioanalysis 13(1): 13-28. PMID: 33319585. PMCID: PMC7739400.

33. Morley GL, Taylor S, Jossi S, Perez-Toledo M, Faustini SE, Marcial-Juarez E, Shields AM, Goodall M, Allen JD, Watanabe Y, Newby ML, Crispin M, Drayson MT, Cunningham AF, Richter AG, O’Shea MK (2020). Sensitive Detection of SARS-CoV-2-Specific Antibodies in Dried Blood Spot Samples. Emerg Infect Dis 26(12): 2970-3. PMID: 32969788. PMCID: PMC7706975.

34. McDade TW, McNally EM, Zelikovich AS, D’Aquila R, Mustanski B, Miller A, Vaught LA, Reiser NL, Bogdanovic E, Fallon KS, Demonbreun AR (2020). High seroprevalence for SARS-CoV-2 among household members of essential workers detected using a dried blood spot assay. PLoS One 15(8): e0237833. PMID: 32797108. PMCID: PMC7428174 competing interests: Thomas W. McDade has a financial interest in EnMed Microanalytics, a company that performs laboratory tests using dried blood spot samples. There are no patents, products in development or marketed products associated with this research to declare. This does not alter our adherence to PLOS ONE policies on sharing data and materials.

35. Turgeon CT, Sanders KA, Granger D, Nett SL, Hilgart H, Matern D, Theel ES (2021). Detection of SARS-CoV-2 IgG antibodies in dried blood spots. Diagn Microbiol Infect Dis 101(1): 115425. PMID: 34116343. PMCID: PMC8116316.

36. Turgeon CT, Sanders KA, Rinaldo P, Granger D, Hilgart H, Matern D, Theel ES (2021). Validation of a multiplex flow immunoassay for detection of IgG antibodies against SARS-CoV-2 in dried blood spots. PLoS One 16(5): e0252621. PMID: 34048503. PMCID: PMC8162624 Theel has the following broad interests that could be perceived as conflicts of interest: - Consulting fees from Roche Diagnostics and Accelerate Diagnostics - Member of the College of American Pathologists’ Microbiology Resource Committee (travel compensation) - Editor, Clinical Microbiology Newsletter This does not alter our adherence to PLOS ONE policies on sharing data and materials.

37. Amanat F, Stadlbauer D, Strohmeier S, Nguyen THO, Chromikova V, McMahon M, Jiang K, Arunkumar GA, Jurczyszak D, Polanco J, Bermudez-Gonzalez M, Kleiner G, Aydillo T, Miorin L, Fierer DS, Lugo LA, Kojic EM, Stoever J, Liu STH, Cunningham-Rundles C, Felgner PL, Moran T, Garcia-Sastre A, Caplivski D, Cheng AC, Kedzierska K, Vapalahti O, Hepojoki JM, Simon V, Krammer F (2020). A serological assay to detect SARS-CoV-2 seroconversion in humans. Nat Med 26(7): 1033-6. PMID: 32398876. PMCID: PMC8183627.

38. Stadlbauer D, Amanat F, Chromikova V, Jiang K, Strohmeier S, Arunkumar GA, Tan J, Bhavsar D, Capuano C, Kirkpatrick E, Meade P, Brito RN, Teo C, McMahon M, Simon V, Krammer F (2020). SARS-CoV-2 Seroconversion in Humans: A Detailed Protocol for a Serological Assay, Antigen Production, and Test Setup. Curr Protoc Microbiol 57(1): e100. PMID: 32302069. PMCID: PMC7235504.

39. Stoddard CI, Galloway J, Chu HY, Shipley MM, Sung K, Itell HL, Wolf CR, Logue JK, Magedson A, Garrett ME, Crawford KHD, Laserson U, Matsen FAt, Overbaugh J (2021). Epitope profiling reveals binding signatures of SARS-CoV-2 immune response in natural infection and cross-reactivity with endemic human CoVs. Cell Rep 35(8): 109164. PMID: 33991511. PMCID: PMC8121454.

40. Crawford KHD, Dingens AS, Eguia R, Wolf CR, Wilcox N, Logue JK, Shuey K, Casto AM, Fiala B, Wrenn S, Pettie D, King NP, Greninger AL, Chu HY, Bloom JD (2020). Dynamics of neutralizing antibody titers in the months after SARS-CoV-2 infection. J Infect Dis. PMID: 33000143.

41. Addetia A, Crawford KHD, Dingens A, Zhu H, Roychoudhury P, Huang ML, Jerome KR, Bloom JD, Greninger AL (2020). Neutralizing Antibodies Correlate with Protection from SARS-CoV-2 in Humans during a Fishery Vessel Outbreak with a High Attack Rate. J Clin Microbiol 58(11). PMID: 32826322. PMCID: PMC7587101.

42. Daag JV, Ylade M, Jadi R, Adams C, Cuachin AM, Alpay R, Aportadera ETC, Yoon IK, de Silva AM, Lopez AL, Deen J (2021). Performance of Dried Blood Spots Compared with Serum Samples for Measuring Dengue Seroprevalence in a Cohort of Children in Cebu, Philippines. Am J Trop Med Hyg 104(1): 130-5. PMID: 33146119. PMCID: PMC7790110.

43. Fachiroh J, Prasetyanti PR, Paramita DK, Prasetyawati AT, Anggrahini DW, Haryana SM, Middeldorp JM (2008). Dried-blood sampling for epstein-barr virus immunoglobulin G (IgG) and IgA serology in nasopharyngeal carcinoma screening. J Clin Microbiol 46(4): 1374-80. PMID: 18256216. PMCID: PMC2292939.

44. Stefic K, Guinard J, Peytavin G, Saboni L, Sommen C, Sauvage C, Lot F, Laperche S, Velter A, Barin F (2019). Screening for Human Immunodeficiency Virus Infection by Use of a Fourth-Generation Antigen/Antibody Assay and Dried Blood Spots: In-Depth Analysis of Sensitivity and Performance Assessment in a Cross-Sectional Study. J Clin Microbiol 58(1). PMID: 31666365. PMCID: PMC6935938.

45. Mohan D, Wansley DL, Sie BM, Noon MS, Baer AN, Laserson U, Larman HB (2018). PhIP-Seq characterization of serum antibodies using oligonucleotide-encoded peptidomes. Nat Protoc 13(9): 1958-78. PMID: 30190553. PMCID: PMC6568263.

46. Dingens AS, Crawford KHD, Adler A, Steele SL, Lacombe K, Eguia R, Amanat F, Walls AC, Wolf CR, Murphy M, Pettie D, Carter L, Qin X, King NP, Veesler D, Krammer F, Dickerson JA, Chu HY, Englund JA, Bloom JD (2020). Serological identification of SARS-CoV-2 infections among children visiting a hospital during the initial Seattle outbreak. Nat Commun 11(1): 4378. PMID: 32873791. PMCID: PMC7463158.

47. Williams KL, Stumpf M, Naiman NE, Ding S, Garrett M, Gobillot T, Vezina D, Dusenbury K, Ramadoss NS, Basom R, Kim PS, Finzi A, Overbaugh J (2019). Identification of HIV gp41-specific antibodies that mediate killing of infected cells. PLoS Pathog 15(2): e1007572. PMID: 30779811. PMCID: PMC6396944.

